# Prediction of Postoperative Delirium in Older Adults from Preoperative Cognition and Occipital Alpha Power from Resting-State Electroencephalogram

**DOI:** 10.1101/2024.08.15.24312053

**Authors:** Matthew Ning, Andrei Rodionov, Jessica M. Ross, Recep A. Ozdemir, Maja Burch, Shu Jing Lian, David Alsop, Michele Cavallari, Bradford C. Dickerson, Tamara G. Fong, Richard N. Jones, Towia A. Libermann, Edward R. Marcantonio, Emiliano Santarnecchi, Eva M. Schmitt, Alexandra Touroutoglou, Thomas G. Travison, Leah Acker, Melody Reese, Haoqi Sun, Brandon Westover, Miles Berger, Alvaro Pascual-Leone, Sharon K. Inouye, Mouhsin M. Shafi, the SAGES II Study Group and the INTUIT/PRIME Study Groups

## Abstract

**Background:** Postoperative delirium is the most common complication following surgery among older adults, and has been consistently associated with increased mortality and morbidity, cognitive decline, and loss of independence, as well as markedly increased health-care costs. Electroencephalography (EEG) spectral slowing has frequently been observed during episodes of delirium, whereas intraoperative frontal alpha power is associated with postoperative delirium. We sought to identify preoperative predictors that could identify individuals at high risk for postoperative delirium, which could guide clinical decision-making and enable targeted interventions to potentially decrease delirium incidence and postoperative delirium-related complications.

**Methods:** In this prospective observational study, we used machine learning to evaluate whether baseline (preoperative) cognitive function and resting-state EEG could be used to identify patients at risk for postoperative delirium. Preoperative resting-state EEGs and the Montreal Cognitive Assessment were collected from 85 patients (age = 73 ± 6.4 years, 12 cases of delirium) undergoing elective surgery. The model with the highest f1-score was subsequently validated in an independent, prospective cohort of 51 older adults (age = 68 ± 5.2 years, 6 cases of delirium) undergoing elective surgery.

**Results:** Occipital alpha powers have higher f1-score than frontal alpha powers and EEG spectral slowing in the training cohort. Occipital alpha powers were able to predict postoperative delirium with AUC, specificity and accuracy all >90%, and sensitivity >80%, in the validation cohort. Notably, models incorporating transformed alpha powers and cognitive scores outperformed models incorporating occipital alpha powers alone or cognitive scores alone.

**Conclusions:** While requiring prospective validation in larger cohorts, these results suggest that strong prediction of postoperative delirium may be feasible in clinical settings using simple and widely available clinical tools. Additionally, our results suggested that the thalamocortical circuit exhibits different EEG patterns under different stressors, with occipital alpha powers potentially reflecting baseline vulnerabilities.

**Clinical Trials:**

1. INTUIT: Investigating Neuroinflammation Underlying Postoperative Cognitive Dysfunction (ClinicalTrials.gov ID: NCT03273335, PI: Miles Berger, Project Start Date: 2017-06-15)

**Prior Presentation:** 2024 American Delirium Society Annual Conference, presented by Matthew Ning, Ph.D., June 11^th^, 2024, Sacramento, CA, USA.

**Preprint Server:** URL: https://www.medrxiv.org/content/10.1101/2024.08.15.24312053v1

## Introduction

Delirium is a complex neuropsychiatric syndrome that is characterized by an acute, fluctuating disturbance in attention, level of consciousness, and cognition.^1^ Postoperative delirium is a common complication in older adults after surgery, occurring in 19-32% of patients,^2^ and is associated with longer ICU and hospital stay, increased post-discharge institutionalization, persistent cognitive decline, and increased short- and long-term mortality.^3–7^ Postoperative delirium has an estimated annual healthcare cost of over $30 billion in the USA alone.^8^

Early and accurate identification of individuals at high risk could enable interventions to reduce the incidence, severity and duration of postoperative delirium. Such interventions might include a careful evaluation of the risk-benefit ratio for surgery; enabling brain health optimization prior to surgery e.g. via elimination of medications that increase delirium risk or through pre-operative transcranial direct current stimulation;^9^ modification of intraoperative anesthesia;^10^ postoperative treatment with acetaminophen;^11^ and through targeted implementation of more intensive behavioral protocols before and after the surgery (e.g. the ABCDEF Bundle,^12^ HELP^13, 14^ and mHELP programs,^15^ where the latter achieved a 56% reduction of postoperative delirium incidence rate within the intervention versus control group). Many studies have shown that preoperative cognitive impairment is a strong predictor of postoperative delirium.^16, 17^ Other risk factors include age, history of alcohol abuse, history of smoking, medical comorbidity and pre-existing impairment in activities of daily living.^18, 19^ However, while these risk factors increase delirium risk at the group level, there are limited clinically useful tools to predict delirium at the individual level. Furthermore, despite studies linking abnormalities in cerebral oscillatory activity with delirium,^20^ the specific mechanisms by which risk factors for delirium are related to the underlying brain dysfunction and subsequent delirium symptoms are still not fully understood.

While the etiology of postoperative delirium is commonly accepted to be multifactorial,^18^ one ongoing theory is that baseline vulnerabilities or cognitive functions may play a larger role than anesthetic or sedative depth in postoperative delirium.^21^ In line with this theory, we hypothesized a neurophysiological model of delirium,^22^ where delirium is the result of impaired cognitive functions in individuals with pre-existing impairments in brain connectivity and plasticity exposed to a stressor, such as surgery. EEG is a neuroimaging technology to measure cortical connectivity and physiology. EEG slowing, a shift in EEG spectral power from high frequency to low-frequency, is frequently seen in subjects with delirium.^23–25^ In addition, intraoperative EEG metrics such as frontal alpha powers are associated with baseline cognitive impairment^20, 26^ and up to a 4 fold increase in delirium risk.^27, 28^ More recently, work by ourselves and others has also found preliminary evidence of association between preoperative resting-state EEG (rsEEG) power ratios and postoperative delirium.^29, 30^ However, both preoperative studies were limited to individuals without preoperative cognitive impairments.

Based on our conceptual model of delirium^22^ and the studies described above, we hypothesized that classical machine learning applied to preoperative resting-state EEG features and baseline measures of cognitive function can predict individual postoperative delirium risk. We evaluated a range of models in one prospective cohort of older adults undergoing elective surgery, and the best-performing model was subsequently validated in a second independent prospective cohort of older adults undergoing surgery at another institution. We also assessed whether models combining EEG and cognitive function performed better than those using either EEG or cognitive testing alone.

## Materials and Method

### Participants

The dataset from Successful Aging after Elective Surgery renewal (SAGES II, NIH-NIA P01AG031720) study,^31^ a prospective observational cohort study of older adults scheduled for major elective non-cardiac surgery, was used for the model selection process. SAGES II here will be referred to as SAGES hereafter for brevity. In that study, 420 participants were enrolled. Inclusion criteria were age ≥60 years, English speakers, ability to communicate verbally, scheduled for elective surgery at the following Harvard-affiliated hospitals: Beth Israel Deaconess Medical Center (BIDMC), Brigham and Women’s Hospital (BWH), or Brigham and Women’s Faulkner Hospital (BWFH), and availability for in-person interviews. Exclusion criteria were delirium at baseline, prior hospitalization within 3 months, current hemodialysis or cancer chemotherapy, legal blindness, severe deafness, terminal condition and history of heavy alcohol abuse or withdrawal. Of the total SAGES cohort of 420 participants, a subgroup (*n*=92), meeting additional criteria (Supplementary material), participated in the EEG study. Of these, two (2.2%) participants were dropped from the EEG study due to a surgery cancellation. The remaining 90 participants (age = 72.5±6.3, range 65-93; 33 males, 57 females) were included in the study sample. Five participants were excluded due to low EEG data quality. Thus, 85 participants remain available for the data analysis. Of these, 12 participants (14% of the analysis sample) developed delirium. Demographic and clinical information for the analytic sample is presented in Table 1 (see Supplementary Table S1 for the excluded cohort). Written informed consent for study participation was obtained from all participants according to procedures approved by the institutional review boards of Beth Israel Deaconess Medical Center, Brigham and Women’s Hospital, and the Brigham and Women’s Faulkner Hospital — the study hospitals, and Hebrew SeniorLife — the study coordinating center, all located in Boston, Massachusetts.

The dataset used for model validation is from Investigating Neuroinflammation Underlying Postoperative Cognitive Dysfunction (INTUIT),^32^ also a prospective observational cohort study registered on clinicaltrials.gov (NCT03273335). INTUIT enrolled 201 participants undergoing non-cardiac/non-neurologic surgeries lasting ≥2 hours with a planned postoperative overnight hospitalization. Inclusion criteria were age ≥60 years, English speakers, ability to communicate verbally and scheduled for non-cardiac/non-neurologic surgery at Duke University Medical Center or Duke Regional Hospital, and available for in-person study visits. Unlike SAGES, INTUIT had no exclusion criteria based on preoperative cognition or delirium status, however, none of the subjects had delirium prior to their scheduled surgery. Exclusion criteria were 1) patients on immunosuppressant (e.g. steroids) or immunomodulatory therapy, chemotherapeutic agents with known cognitive effects, or anticoagulants that would preclude safe lumbar puncture, 2) inmates of correctional facilities, 3) patients who experienced major head trauma or received chemotherapy between the baseline and either postoperative cognitive testing session. A subgroup of INTUIT study participants (n=81) participated in an EEG study funded by the separate PRIME study grant.^33^ 63 out of the 81 INTUIT/PRIME EEG participants wore a 32-channel EEG cap with standard international 10-20 montage (Supplementary Figure S1A). The remaining 15 subjects wore a custom 32-channel EEG cap (Supplementary Figure S1B). Due to the lack of overlapping channels with the standard international 10-20 montage, subjects with the custom EEG montage were excluded from further analysis. 1 participant withdrew from the study. EEG data was unable to be collected from 3 participants and 11 participants’ EEG data was excluded due to poor EEG quality, leaving 51 INTUIT participants with EEG data available for analysis, of whom 6 developed delirium (12% of the analysis sample). Demographic and clinical information for this 51 patient cohort from the INTUIT/PRIME study is presented in Table 2. Written informed consent was obtained from all participants. The study was approved by Duke University Health System Institutional Review Board.

### Clinical Assessments

For the SAGES Study, delirium was assessed with the Confusion Assessment Method (CAM) long-form daily post-surgery. CAM is a standardized and internationally accepted tool that enables non-psychiatrically trained clinicians to identify and recognize delirium quickly and accurately in both clinical and research settings (sensitivity 94-100%, specificity 90-95%, interrater reliability 84-100%).^34^ Baseline cognitive function was assessed with The Montréal Cognitive Assessment (MoCA, range 0-30, 0=most impaired).^35^ MoCA were conducted either through video call over Zoom or face-to-face in the patient’s place of residence prior to surgery. A MoCA score of 26 or above was considered normal while scores less than 26 were considered as indicative of possible cognitive impairment. Because the MoCA was administered after other neuropsychological assessments, the memory subdomain test of MoCA was corrected for possible interference effect from the previous memory tests (See Supplementary materials). For the INTUIT/PRIME Study, a short form of CAM, called the 3D-CAM, was used to identify delirium. The 3D-CAM has excellent overall agreement with the long-form CAM.^36, 37^ The Mini-Mental Status Examination (MMSE) was used in the INTUIT/PRIME study for assessment of baseline cognitive function (range 0-30, 0-most impaired). The MMSE was converted to MoCA scores based on an established cross-walk.^38, 39^ Since one MMSE score can be mapped to multiple MoCA scores, the median of multiple MoCA scores was used for our study following recommended conversion procedures (see Supplementary Table 3 for the conversion table).

### EEG Data Collection

For the SAGES EEG Sub-Study, participants attended a baseline EEG visit at least 2-3 days before and up to two months in advance of their scheduled surgery. RsEEG was recorded at 5 kHz while participants were seated in a comfortable chair during a 3-4 hour study visit that included collection of Transcranial Magnetic Stimulation and EEG data. RsEEG was the first neurophysiological measurement taken during the study visit and was recorded for 3-7 minutes each in two conditions: with eyes open (EO) and eyes closed (EC). For the eyes closed condition, the participants were instructed to maintain wakefulness, and were queried intermittently to ensure alertness. RsEEG was recorded with a 64-channel Brainproducts system with AFz as the reference channel and the electrode impedances < 20 kΩ (extended 10-20 international system, reference channel: AFz, Supplementary Figure S1C) and amplifiers (actiCHamp, Brain Products GmbH, Munich, Germany). All participants tolerated the procedure and no significant side effects were reported or noticed.

For the INTUIT/PRIME Study, rsEEG was recorded at 1 kHz from 32 electrodes embedded in a 64-electrode cap custom designed for extended scalp coverage,^38^ with Cz as the reference channel and the electrode impedances < 20 kΩ. (BrainAmpMR Plus, Brain Products GmbH, Gilching, Germany). INTUIT/PRIME rsEEG was recorded in the preoperative patient holding area just prior to surgery, for 3 min each with eyes open and with eyes closed. Participants were also instructed and monitored to maintain wakefulness during the eyes closed condition.

### EEG Processing and Analysis

EEG processing details are in the Supplementary Method section.

### Features and Transformation

EEG spectral power ratio (SPR), defined as (alpha+beta power)/(delta+theta power), as well as frontal alpha powers, were hypothesized a priori to predict postoperative delirium. SPRs from all 59 channels in both eye conditions, plus MoCA, make up the SPR Feature Set. Alpha powers from Fp1, Fp2, F7 and F8 channels in both eye conditions, plus MoCA, make up the Frontal Alpha Powers Set. In addition, for the preliminary data exploration on the SAGES cohort (Figure 1A), we systematically examined EEG spectral powers of several different frequency bands as well as ratios of frequency band powers from different regions of interest (ROIs). Through this process, alpha powers (8-12 Hz) from the occipital region were identified as promising candidates for further analysis and model development. The feature set with the transformed occipital alpha powers and MoCA will be referred to throughout this paper as the Principal Feature Set. More information is in the Methods section of the Supplementary Materials.

**Figure 1.**
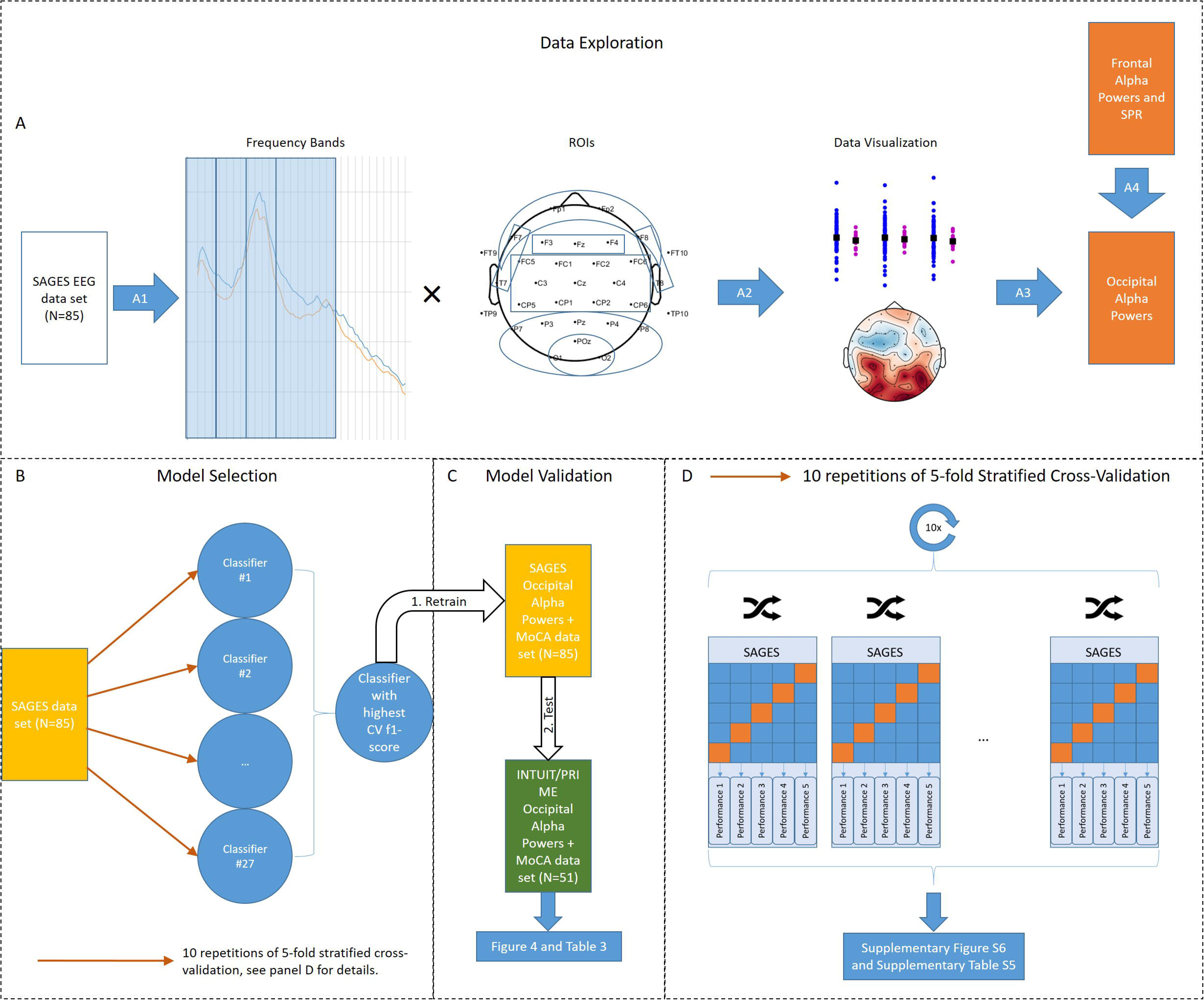
Schematic diagram of machine learning framework. A) Data exploration. A1) different band powers of different regions of interested (ROIs) were extracted from SAGES EEG Data Set. A2) the results are visualized using scatter plots as well as EEG topographic plots. A3) occipital alpha powers (and sub-alpha powers) from both eyes-open and eyes-closed condition was selected. Baseline cognition (MoCA) was selected a priori. B) Model Selection. The feature sets consisting of EEG alpha (and sub-alpha) powers determined from data exploration (A), along with MoCA selected a priori, were tested using 9 different classifiers using cross-validation (red arrows). The details of the cross-validation is shown in (D). C) After the classifier with the highest f1-score is determined in model selection step, the classifier was re-trained on the entire SAGES Data Set, then the parameters of the classifier were held fixed and independently validated on INTUIT/PRIME Data Set. The results are plotted in Figure 4 and tabulated in Table 3. D) Details of the cross-validation used in model selection step. The CV performance are plotted in Supplementary Figure S6 and Supplementary Table S6.

### ML Models and Cross-Validation

The SAGES cohort was used for the model selection step (Figure 1B). The following models were tested during the model selection step: logistic regression with L2 regularization with inner cross-validation (CV) for hyperparameter tuning; linear discriminant analysis (LDA) with inner CV for hyperparameter tuning; linear discriminant analysis (LDA) using Ledoit-Wolf estimator (LDA LW); LDA using Oracle Shrinkage Approximation estimator (LDA OA); nearest shrunken centroid (NSC) using Manhattan distance metric; NSC using Euclidean distance metric; Gaussian Naïve Bayes (GNB) using empirical priors; GNB using a priori-defined class priors (80-20 ratio) and decision tree. During the model selection step, the SAGES Data Set was cross-validated using 10 repetitions of 5-fold stratified CV (Figure 1D). The 95% confidence intervals were estimated using the student’s t-distribution using the sample mean and standard deviation of 50 folds. The models were assessed with 8 metrics: accuracy, sensitivity, specificity, f1-score, area under the curve of the receiver operating characteristic curve (ROC-AUC), precision-recall curve (PR-AUC) and positive predictive value (PPV, also known as precision) and negative predictive value (NPV). To prioritize the minority class (delirium), f1-score was chosen as the deciding factor. The INTUIT/PRIME cohort was used for the model validation (Figure 1C). For the model validation step, the best performing model from the model selection step was chosen based on f1-score, retrained on the entire SAGES Data Set, and model parameters were then fixed and tested on the entire INTUIT/PRIME Data Set. Bootstrapping was used to compute the 95% confidence intervals, where the test set (INTUIT/PRIME Data Set) was re-sampled 2000 times, with each re-sampling set stratified to the class proportions of the original sample.

## Result

Scatterplots of the untransformed (left panel) and transformed (right panel) resting state alpha powers from the O2 channel are shown in Figure 2. In the untransformed samples, the control group (blue) has similar means but much larger variances than the delirium group (purple) in both data sets and eye conditions, with no significant differences between groups. The variances of both SAGES and INTUIT/PRIME samples are remarkably similar within the same eye conditions and groups. After transformation, all of the group contrasts are significant (two-sample two-tailed Welch’s t-test, α<0.05). Findings are similar for the resting state alpha powers in the other channels in the occipital region (Supplementary Figure S2 and S3). The PSD plots for the O2 channel in both SAGES and INTUIT/PRIME Data Sets are shown in Figure 3. While, the alpha peaks are consistent across both data sets within the same eye conditions and groups, the distributions of the powers within the delta, theta and beta frequency ranges are not consistent. Similar findings arise in O1 and POz channels (Supplementary Figure S4 and S5, respectively).

**Figure 2.**
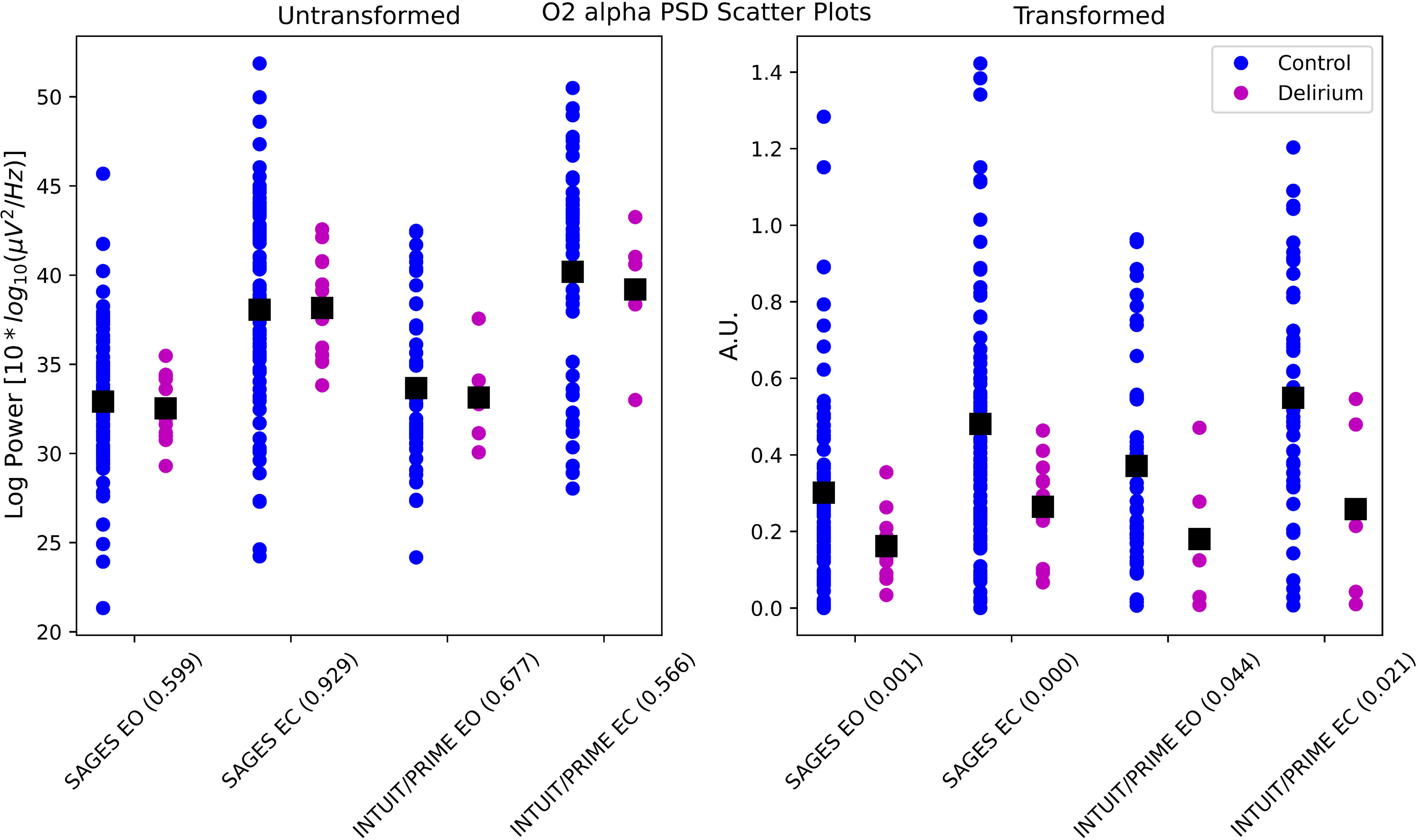
Scatter plots of individual EEG alpha powers in O2 channel. Scatter plot showing distributions of alpha powers in participants with post-operative delirium (purple dots) and without delirium (blue dots). Black squares represent the mean of their respective groups. Left panel shows the untransformed alpha powers as seen in PSD plots (Fig. 3) and right panel shows transformed alpha powers. Values enclosed in parentheses in the x axis represent the p-values of two-sample two-tailed Welch’s t-test. This channel is representative of all channels in the occipital region and their scatter plots can be found in the Supplementary Fig. S2 & S3.

**Figure 3.**
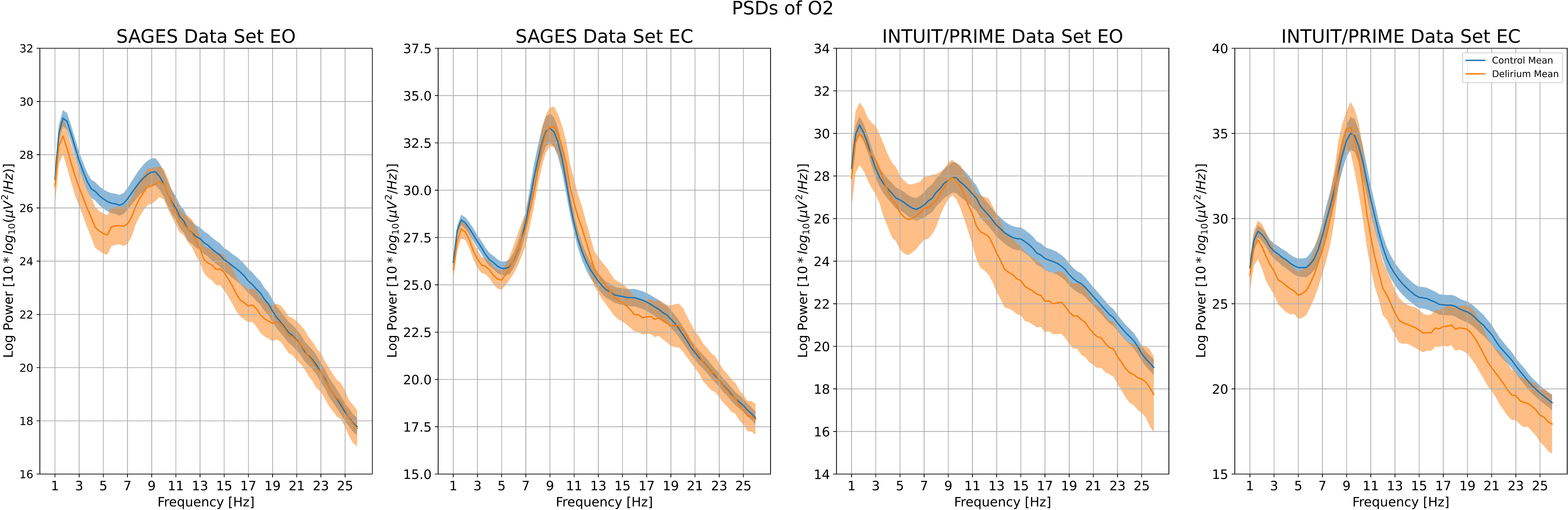
Power spectral densities of O2 channel. Power spectral densities of O2 channel from both SAGES and INTUIT/PRIME Data Sets in both eyes-open (EO) and eyes-closed (EC) conditions for [1, 26] Hz frequency range. Shaded regions represent standard error of the means. Blue represents control group and orange represents delirium group. PSDs for O1 and POz are shown in Supplementary Figure S4 & S5, respectively.

The cross-validation results of SAGES Data Set of the SPR Feature Set, Frontal Alpha Powers Set and the Principal Feature Set are shown in Supplementary Figure S6 and Supplementary Table S5. Out of the 27 models tested, LDA LW model based on the Principal Feature Set has the highest f1-score (mean ± 95% confidence interval: 0.57 ± 0.07, sensitivity: 0.54 ± 0.08, specificity: 0.94 ± 0.02, ROC-AUC: 0.80 ± 0.04). With the Principal Feature Set selected, when retraining and then testing the LDA LW model on the entire INTUIT/PRIME Data Set (Supplementary Table S5), the f1-score is 0.67 (95% CI [0.43, 0.92]), sensitivity and specificity are 0.83 (95% CI [0.50, 1.00]) and 0.91 (95% CI [0.82, 0.98]), respectively. The ROC and precision-recall curves are shown in Figure 4B-C. Their AUCs are 0.94, 95% CI [0.87, 0.99] and 0.70, 95% CI [0.46, 0.95], respectively. When compared to MoCA-Alone (Table 3), the Principal Feature Set outperformed MoCA alone in all metrics except for sensitivity (tied at 0.83).

**Figure 4.**
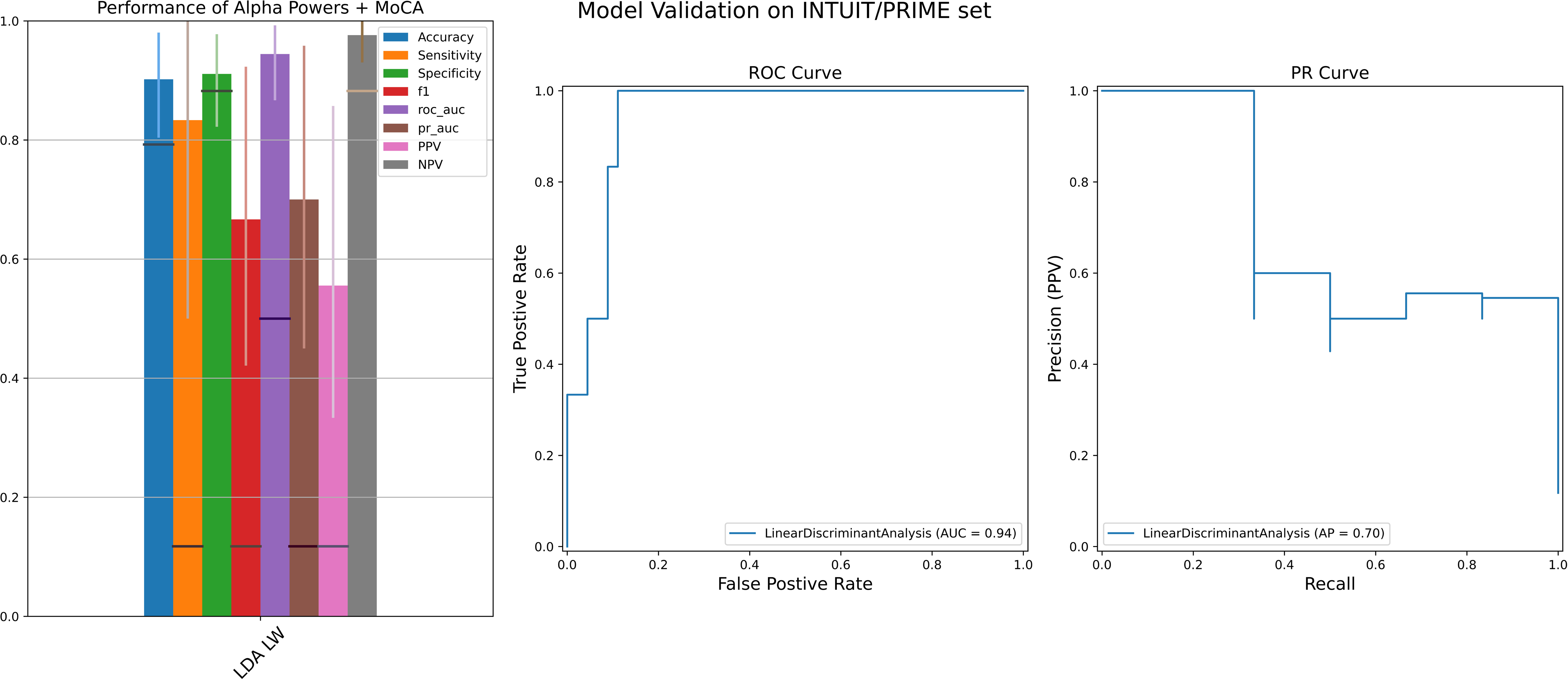
Model validation: performances using the Principal Feature Set (Alpha Powers + MoCA). A) The performance of the model (LDA LW) selected from the model selection step. 95% confidence intervals are shown as thin vertical bars. Chance levels for each metric are shown as dark horizontal lines (details of chance levels in Supplementary Materials). Blue: accuracy, orange: sensitivity, green: specificity, red: f1 score, purple: AUC of ROC curve, brown: AUC of precision-recall curve, pink: positive predictive value (PPV, also known as precision), grey: negative predictive value (NPV). Values are tabulated in Table 4A. ROC (B) and PR (C) curves are shown for the selected LDA LW model. The AUCs reported as mean and 95% CI are 0.94 [0.86, 0.99] for the ROC curve and 0.70 [0.44, 0.95] for the PR curve.

MoCA-Alone yielded sensitivity, specificity and ROC-AUC of 0.83 (95% CI [0.50, 1.00]), 0.80 (95% CI [0.67, 0.91]), 0.91 (95% CI [0.81, 0.98]) respectively. The biggest improvements from MoCA-Alone to the Principal Feature Set lie in PPV (0.20 increase) and specificity (0.11 increase). When compared to Alpha-Powers-Alone (Table 3), Alpha-Powers-Alone has very skewed performance between sensitivity (0.17, 95% CI [0, 0.50]) and specificity (1.00, 95% CI [1.00, 1.00]). Alpha-Powers-Alone’s ROC-AUC is 0.77, 95% CI [0.57, 0.94].

## Discussion

Using preoperative rsEEG and baseline cognitive functions collected from 2 independent cohorts of older adults undergoing elective surgeries, we trained and tested machine learning models to assess their performance in predicting post-operative delirium. In the model selection step, the Principal Feature Set, consisting of resting state EEG occipital alpha powers and baseline MoCA scores outperformed the SPR Feature Set and the Frontal Alpha Powers Set in the stratified cross-validation of the SAGES Data Set.

Using the Principal Feature Set, LDA LW, the model with the highest f1-score was selected and retrained on the entire SAGES Data Set and with the model parameters fixed, tested on an independently collected cohort, the INTUIT/PRIME Data Set. In the INTUIT/PRIME Data Set, LDA LW yielded ≥ 0.9 in accuracy, specificity and ROC-AUC and ≥ 0.8 in sensitivity. Importantly, it substantially outperformed MoCA-Alone, with the biggest improvements in PPV (0.20 increase) and specificity (0.11 increase). This finding has two major implications: 1) rsEEG can be used in conjunction with the MoCA to preoperatively screen for patients at high risk for post-operative delirium, and 2) occipital alpha powers represent a candidate risk factor that can be studied further to understand the possible neurobiological etiology of delirium.^40^ Additionally, our results provide support for the theory that baseline vulnerability may play a major role in postoperative delirium and that occipital alpha powers are associated with baseline vulnerability.^21, 22^

The strong predictive performance seen above can be attributed to the difference in the variances of the resting state occipital alpha powers. One possible interpretation of the differences in the variances is that individuals within the control group exhibit greater fluctuation in the EEG powers over time relative to the delirium group and the greater fluctuation could be an indirect indication of cortical connectivity and plasticity.^41–43^ Thus, greater fluctuation over time from one healthy individual could be translated to greater fluctuation across multiple healthy individuals from a fixed time point, relative to individuals with postoperative delirium. While the variances of alpha power over time within one resting state recording session didn’t show strong predictive performance for delirium (results not shown), we hypothesized that the variance over much longer range of time might better measure the integrity of cortical circuits. This could be tested with a longitudinal study with repeated measurements from the same individuals, analogous to precision functional mapping. Alternatively, exploring functional connectivity could reveal more direct information about the cortical connectivity.^41^

There are only a few studies on the relationship between preoperative EEG and postoperative delirium and all of them used regressions/statistical tests to establish association. Kim et al.^30^ used logistic regression to show association between the median dominant frequency of the prefrontal cortical region (Fp1 and Fp2) and postoperative delirium. Similarly, while Schuβler et al.^44^ recorded EEG using 10-20 international system, they limited their report to F1 and F2 channels. They used statistical tests to show a significant decrease in the power of the high beta and low gamma bands. One study that was done on the same INTUIT/PRIME dataset used here, but analyzed independently of this study, used multivariate logistic and proportion-odds regression analysis to show that preoperative alpha power attenuation, the reduction in alpha power from eyes-closed to eyes-opened condition, is inversely associated with postoperative attention^37^. However, based on our internal testing, the alpha power attenuations, either taken as the difference or ratio between eyes-closed and eyes-opened conditions, don’t have the same predictive performance of delirium (not shown) as our proposed model. In contrast, one recurring and consistent finding of EEG during episodes of delirium is the EEG slowing.^28, 45–48^ Another consistent finding in the intraoperative setting is the significant association between lower frontal alpha powers and postoperative delirium.^28^ However, in the preoperative setting, both EEG slowing, as indexed by spectral power ratio, and frontal alpha powers do not have the same predictive performance as occipital alpha powers. The connection between EEG slowing during delirium, lower frontal alpha powers during surgeries and restricted range of occipital alpha powers observed preoperatively here is unclear, but one possibility is that the thalamocortical connectivity responsible for the generation of the alpha rhythm is particularly susceptible to different breakdowns during different stressors. If true, this implies that thalamocortical connectivity is a measure of baseline vulnerability, which, in turn, plays a bigger role in postoperative delirium than other factors and would explain the failure of studies targeting depth-of-anesthesia to reduce the risk of post-operative delirium.^49^ More research is needed to better understand the connection.

Improvement in predictive performance of postoperative delirium over pre-operative cognitive assessment alone is one of the major findings in this study. Past studies on pre-operative prediction of postoperative delirium have largely focused on different measures of cognitive functioning, including MoCA.^16, 17^ While lower cognitive scores have been independently associated with higher risk of developing postoperative delirium, the relationship and especially the pathophysiology and etiology between cognitive functioning and postoperative delirium are still not fully understood. Additionally, cognitive functioning is not the sole risk factor for postoperative delirium.^50^ Instead, risk is assumed to be multifactorial but is primarily determined by both patient-related predisposing factors (which include cognitive impairment) and treatment-associated precipitating factors.^18, 19^ Our analysis suggests that incorporation of neurophysiological features such as resting-state EEG measures improve model performance. This in turn provides support for our conceptual model linking postoperative delirium to baseline vulnerabilities, and more specifically underlying changes in cerebral physiology.^21, 22^ Lastly, whether the occipital alpha powers is connected solely to baseline cognition or to different precursors for postoperative delirium is unclear.

One main limitation of this study is the difference in the predictive performance in the SAGES and INTUIT/PRIME cohorts, especially in the sensitivity metric. One possible explanation is that the rsEEG for the INTUIT/PRIME cohort was recorded on the same day of the surgery, whereas the rsEEG for the SAGES cohort was recorded at least 2-3 days and up to 2 months in advance of scheduled surgeries. Another main limitation is the relatively small sizes of the training set (SAGES) and especially the test set (INTUIT/PRIME). The confidence intervals of sensitivity and precision are wide (Table 3). Thus, a larger sample size is needed to confirm our findings.

In conclusion, machine learning techniques utilizing preoperative rsEEG and MoCA performance features can predict postoperative delirium with strong performance across multiple metrics. These findings suggest that pre-operative rsEEG can identify subclinical neurophysiological changes that may play a crucial role in the neuropathology of postoperative delirium. Independent confirmation in a larger sample size and with prospective application is needed to validate our proposals, and to potentially improve model performance by incorporation of features from more frequencies and brain regions. If successful, translation into clinical setting would be convenient and straightforward as EEG is a widely available, inexpensive, and well-tolerated neuroimaging technology, and assessment of cognitive performance using the MoCA is relatively fast and simple. The application of such ML approaches to preoperative data thus has the potential to identify individuals at high risk of postoperative delirium, enabling testing of targeted interventions to reduce the risk and morbidity of this common problem.

## Data Availability

The pre-processed and de-identified EEG data is anticipated to be available on a public repository in late 2025. The link to the repository for the EEG data will be posted on the Github repository linked in the next section.

## Acknowledgment

The authors gratefully acknowledge the contributions of the patients, family members, nurses, physicians, staff members, and members of the Executive Committee who participated in the Successful Aging after Elective Surgery (SAGES) Study and the patients and family members who participated in the INTUIT/PRIME study.

## Reference

1. American Psychiatric Association, American Psychiatric Association: Diagnostic and statistical manual of mental disorders: DSM-5, 5th ed. Washington, D.C, American Psychiatric Association, 2013

2. Yan E, Veitch M, Saripella A, et al.: Association between postoperative delirium and adverse outcomes in older surgical patients: A systematic review and meta-analysis. J Clin Anesth 2023; 90:111221

3. Dolan MM, Hawkes WG, Zimmerman SI, et al.: Delirium on hospital admission in aged hip fracture patients: prediction of mortality and 2-year functional outcomes. J Gerontol A Biol Sci Med Sci 2000; 55:M527–534

4. Ely EW, Shintani A, Truman B, et al.: Delirium as a predictor of mortality in mechanically ventilated patients in the intensive care unit. JAMA 2004; 291:1753–62

5. Ouimet S, Kavanagh BP, Gottfried SB, Skrobik Y: Incidence, risk factors and consequences of ICU delirium. Intensive Care Med 2007; 33:66–73

6. Ní Chróinín D, Francis N, Wong P, Kim YD, Nham S, D’Amours S: Older trauma patients are at high risk of delirium, especially those with underlying dementia or baseline frailty. Trauma Surg Acute Care Open 2021; 6:e000639

7. Sanchez D, Brennan K, Al Sayfe M, et al.: Frailty, delirium and hospital mortality of older adults admitted to intensive care: the Delirium (Deli) in ICU study. Crit Care 2020; 24:609

8. Gou RY, Hshieh TT, Marcantonio ER, et al.: One-Year Medicare Costs Associated With Delirium in Older Patients Undergoing Major Elective Surgery. JAMA Surg 2021; 156:430–42

9. Tao M, Zhang S, Han Y, et al.: Efficacy of transcranial direct current stimulation on postoperative delirium in elderly patients undergoing lower limb major arthroplasty: A randomized controlled trial. Brain Stimul 2023; 16:88–96

10. Weinstein SM, Poultsides L, Baaklini LR, et al.: Postoperative delirium in total knee and hip arthroplasty patients: a study of perioperative modifiable risk factors. Br J Anaesth 2018; 120:999–1008

11. Subramaniam B, Shankar P, Shaefi S, et al.: Effect of Intravenous Acetaminophen vs Placebo Combined With Propofol or Dexmedetomidine on Postoperative Delirium Among Older Patients Following Cardiac Surgery: The DEXACET Randomized Clinical Trial. JAMA 2019; 321:686–96

12. Marra A, Ely EW, Pandharipande PP, Patel MB: The ABCDEF Bundle in Critical Care. Crit Care Clin 2017; 33:225–43

13. Inouye SK, Bogardus ST, Charpentier PA, et al.: A multicomponent intervention to prevent delirium in hospitalized older patients. N Engl J Med 1999; 340:669–76

14. Inouye SK, Bogardus ST, Baker DI, Leo-Summers L, Cooney LM: The Hospital Elder Life Program: a model of care to prevent cognitive and functional decline in older hospitalized patients. Hospital Elder Life Program. J Am Geriatr Soc 2000; 48:1697–706

15. Chen CC-H, Li H-C, Liang J-T, et al.: Effect of a Modified Hospital Elder Life Program on Delirium and Length of Hospital Stay in Patients Undergoing Abdominal Surgery: A Cluster Randomized Clinical Trial. JAMA Surg 2017; 152:827–34

16. Wong CK, Munster BC van, Hatseras A, et al.: Head-to-head comparison of 14 prediction models for postoperative delirium in elderly non-ICU patients: an external validation study. BMJ Open 2022; 12:e054023

17. Cizginer S, Marcantonio E, Vasunilashorn S, et al.: The Cognitive Reserve Model in the Development of Delirium: The Successful Aging After Elective Surgery Study. J Geriatr Psychiatry Neurol 2017; 30:337–45

18. Inouye SK: Predisposing and precipitating factors for delirium in hospitalized older patients. Dement Geriatr Cogn Disord 1999; 10:393–400

19. Bellelli G, Brathwaite JS, Mazzola P: Delirium: A Marker of Vulnerability in Older People. Front Aging Neurosci 2021; 13:626127

20. Giattino CM, Gardner JE, Sbahi FM, et al.: Intraoperative Frontal Alpha-Band Power Correlates with Preoperative Neurocognitive Function in Older Adults. Front Syst Neurosci 2017; 11:24

21. Pandharipande PP, Whitlock EL, Hughes CG: Baseline Vulnerabilities May Play a Larger Role than Depth of Anesthesia or Sedation in Postoperative Delirium. Anesthesiology 2021; 135:940–2

22. Shafi MM, Santarnecchi E, Fong TG, et al.: Advancing the Neurophysiological Understanding of Delirium. J Am Geriatr Soc 2017; 65:1114–8

23. Jacobson SA, Leuchter AF, Walter DO, Weiner H: Serial quantitative EEG among elderly subjects with delirium. Biol Psychiatry 1993; 34:135–40

24. Zisook S, Braff DL: Delirium: recognition and management in the older patient. Geriatrics 1986; 41:67-67O, 72–3, 77–8

25. Koponen H, Partanen J, Pääkkönen A, Mattila E, Riekkinen PJ: EEG spectral analysis in delirium. J Neurol Neurosurg Psychiatry 1989; 52:980–5

26. Koch S, Feinkohl I, Chakravarty S, et al.: Cognitive Impairment Is Associated with Absolute Intraoperative Frontal α-Band Power but Not with Baseline α-Band Power: A Pilot Study. Dement Geriatr Cogn Disord 2019; 48:83–92

27. Cooter Wright M, Bunning T, Eleswarpu SS, et al.: A Processed Electroencephalogram-Based Brain Anesthetic Resistance Index Is Associated With Postoperative Delirium in Older Adults: A Dual Center Study. Anesth Analg 2022; 134:149–58

28. Gutierrez R, Egaña JI, Saez I, et al.: Intraoperative Low Alpha Power in the Electroencephalogram Is Associated With Postoperative Subsyndromal Delirium. Front Syst Neurosci 2019; 13:56

29. Ross JM, Santarnecchi E, Lian SJ, et al.: Neurophysiologic predictors of individual risk for post-operative delirium after elective surgery. J Am Geriatr Soc 2023; 71:235–44

30. Kim J, Park S, Kim K-N, et al.: Resting-state prefrontal EEG biomarker in correlation with postoperative delirium in elderly patients. Front Aging Neurosci 2023; 15:1224264

31. Ward M, Hshieh TT, Schmitt EM, et al.: Successful aging after elective surgery II: Study cohort description. J Am Geriatr Soc 2024; 72:209–18

32. Berger M, Oyeyemi D, Olurinde MO, et al.: The INTUIT Study: Investigating Neuroinflammation Underlying Postoperative Cognitive Dysfunction. J Am Geriatr Soc 2019; 67:794–8

33. Whitson HE, Crabtree D, Pieper CF, et al.: A template for physical resilience research in older adults: Methods of the PRIME-KNEE study. J Am Geriatr Soc 2021; 69:3232–41

34. Inouye SK, Dyck CH van, Alessi CA, Balkin S, Siegal AP, Horwitz RI: Clarifying confusion: the confusion assessment method. A new method for detection of delirium. Ann Intern Med 1990; 113:941–8

35. Nasreddine ZS, Phillips NA, Bédirian V, et al.: The Montreal Cognitive Assessment, MoCA: a brief screening tool for mild cognitive impairment. J Am Geriatr Soc 2005; 53:695–9

36. Marcantonio ER, Ngo LH, O’Connor M, et al.: 3D-CAM: derivation and validation of a 3-minute diagnostic interview for CAM-defined delirium: a cross-sectional diagnostic test study. Ann Intern Med 2014; 161:554–61

37. Vasunilashorn SM, Devinney MJ, Acker L, et al.: A New Severity Scoring Scale for the 3-Minute Confusion Assessment Method (3D-CAM). J Am Geriatr Soc 2020; 68:1874–6

38. David-Bercholz J, Acker L, Caceres AI, et al.: Conserved YKL-40 changes in mice and humans after postoperative delirium. Brain Behav Immun Health 2022; 26:100555

39. Saczynski JS, Inouye SK, Guess J, et al.: The Montreal Cognitive Assessment: Creating a Crosswalk with the Mini-Mental State Examination. J Am Geriatr Soc 2015; 63:2370–4

40. Acker L, Wong MK, Wright MC, et al.: Preoperative electroencephalographic alpha-power changes with eyes opening are associated with postoperative attention impairment and inattention-related delirium severity. Br J Anaesth 2024; 132:154–63

41. Deco G, Jirsa VK, McIntosh AR: Emerging concepts for the dynamical organization of resting-state activity in the brain. Nat Rev Neurosci 2011; 12:43–56

42. McIntosh AR, Kovacevic N, Itier RJ: Increased brain signal variability accompanies lower behavioral variability in development. PLoS Comput Biol 2008; 4:e1000106

43. Woltering S, Jung J, Liu Z, Tannock R: Resting state EEG oscillatory power differences in ADHD college students and their peers. Behav Brain Funct 2012; 8:60

44. Schüßler J, Ostertag J, Georgii M-T, et al.: Preoperative characterization of baseline EEG recordings for risk stratification of post-anesthesia care unit delirium. J Clin Anesth 2023; 86:111058

45. Palanca BJA, Wildes TS, Ju YS, Ching S, Avidan MS: Electroencephalography and delirium in the postoperative period. Br J Anaesth 2017; 119:294–307

46. Kinoshita H, Saito J, Kushikata T, et al.: The Perioperative Frontal Relative Ratio of the Alpha Power of Electroencephalography for Predicting Postoperative Delirium After Highly Invasive Surgery: A Prospective Observational Study. Anesth Analg 2023; 137:1279–88

47. Guay CS, Kafashan M, Huels ER, et al.: Postoperative Delirium Severity and Recovery Correlate With Electroencephalogram Spectral Features. Anesth Analg 2023; 136:140–51

48. Wiegand TLT, Rémi J, Dimitriadis K: Electroencephalography in delirium assessment: a scoping review. BMC Neurol 2022; 22:86

49. Brown CH, Edwards C, Lin C, et al.: Spinal Anesthesia with Targeted Sedation based on Bispectral Index Values Compared with General Anesthesia with Masked Bispectral Index Values to Reduce Delirium: The SHARP Randomized Controlled Trial. Anesthesiology 2021; 135:992–1003

50. Menzenbach J, Kirfel A, Guttenthaler V, et al.: PRe-Operative Prediction of postoperative DElirium by appropriate SCreening (PROPDESC) development and validation of a pragmatic POD risk screening score based on routine preoperative data. J Clin Anesth 2022; 78:110684

