## Supplementary Text for "Prediction of Postoperative Delirium in Older Adults from Preoperative Cognition and Occipital Alpha Power from Resting-State Electroencephalogram"

Supplementary Materials

Methods

**Clinical assessments**

For the SAGES study, MoCA was conducted either through video call over Zoom or face-to-face in the patient’s place of residence prior to surgery. MoCA was administered after other neuropsychological tests and thus introduced interference in the word recall task. Thus, correction to MoCA scores was applied based on an established method (for details, see Helfand et al^1^.)

**EEG Processing**

EEG processing here is similar to our previous work [24]. In both data sets, EEG signals were first down-sampled to 500 Hz. In order, notch (band stop frequency 57-63 Hz, 4th total order forward-backward Butterworth), high-pass (1 Hz high pass frequency, 4^th^ total order forward-backward Butterworth) and low-pass (50 Hz low-pass frequency, 4th total order forward-backward Butterworth) filters were applied. EEG channels contaminated by artifacts were manually identified and removed (Number of rejected channels reported as average ± SD. SAGES: 2.0 ± 2.0, INTUIT/PRIME: 1.3 ± 1.6. Number of remaining channels reported as average ± SD. SAGES: 60.9 ± 2.0, INTUIT/PRIME: 30.7 ± 1.6). The data were divided into 3-second epochs, visually inspected and bad epochs were manually removed. Number of rejected epochs are reported as average ± SD. SAGES: 2.0 ± 4.7, INTUIT/PRIME: 3.7 ± 4.4. Number of remaining epochs are reported as average ± SD. SAGES: 58.6 ±10.0, INTUIT/PRIME: 56.4 ± 9.6. After that, EEG data were re-referenced to common average reference. Prior to running independent component analysis (ICA) on EEG channels, principal component analysis (PCA) was used to reduce the dimension (30 for SAGES Data Set and 24 for INTUIT/PRIME Data Set). Fast independent component analysis (fICA v2.5, http://research.ics.aalto.fi/ica/fastica/) EEGLAB plugin was used to compute independent components (ICs). Non-brain ICs that represented blink/eye movement, electromyographic activity, single electrode noise, or cardiac beats artifacts were manually identified based on their power spectrum, amplitude, scalp topography, and time course using TMS-EEG Signal Analyser (TESA v1.1.1, <http://nigelrogasch.github.io/TESA>) [36, 37] EEGLAB toolbox (SAGES: Total number of analyzed ICs – 30, average ±SD rejected ICs = 16.1 ±5.0; total average ±SD remaining ICs = 14.0 ±5.0. INTUIT/PRIME: Total number of analyzed ICs – 24, average ±SD rejected ICs = 9.4 ±3.2; total average ±SD remaining ICs = 14.6 ±3.2). Any subjects where more than 85% of the EEG signal variance are removed via ICA were excluded. As a result, for the SAGES Data Set, 5 subjects were excluded from further analysis, leaving 85 subjects available for analysis. For the INTUIT/PRIME Data Set, 12 subjects were excluded from further analysis, leaving 51 subjects remaining. Channels rejected during the previous steps were interpolated using spherical interpolation. Individual channel spectrum plots were inspected and remaining bad channels were removed and interpolated again using spherical interpolation (Number of channel rejected, reported as average ± SD: SAGES: 1.0 ± 1.7, INTUIT/PRIME: 0.2 ± 0.6). The data recorded from the channels FT9, FT10, TP9 and TP10 was contaminated by artifacts in most of the SAGES subjects. To ensure high signal to noise ratio, these channels were removed from all SAGES datasets. These channels weren’t available in the INTUIT/PRIME datasets.

**Features and Transformation**

EEG spectral power ratio, defined as (alpha+beta power)/(delta+theta power), was hypothesized a priori to predict postoperative delirium and is tested here. In addition, for the preliminary data exploration on the SAGES cohort (Figure 1A), we systematically examined EEG spectral powers of several different frequency bands as well as ratios of frequency band powers from different regions of interest (ROIs) to identify features that might potentially predict delirium (Supplementary Table S4). Additionally, we tested two different data transformations: standardization to Gaussian distribution with 0 mean and unit variance, and log of the distance to the median (Equation 1). Power spectral densities (PSDs), scatter plots of band powers and EEG topographic plots all were used. Through this process, alpha (8-12 Hz) and sub-alpha powers (8-10 Hz, 10-12 Hz) from the occipital region were identified as promising candidates for further analysis and model development. For each of 3 electrodes in the occipital region (O1, O2 and POz), the power spectral density (PSD) was estimated using the multitaper method (using Discrete Prolate Spheroidal (Slepian) Sequences as tapers) and was estimated separately for the eyes-open (EO) and eyes-closed (EC) conditions. 3 different sets of features were tested for the classification performance (Supplementary Table S5). For the first set, the z-scores of SPR were computed from each of the 3 electrodes and eye conditions. For the second set, the alpha powers were computed from the PSD for each electrode and condition and were defined as the average powers of the alpha frequency range ([8-12] Hz) and were subsequently transformed to the log of the distance to the median (Equation 1). For the third set, using a bandwidth of 2 Hz, the average band powers were computed using the 8-10 Hz range for the EC condition and the 10-12 Hz range for the EO condition and were transformed to the log of the distance to the median (Equation 1).

$$\tilde{p}_{i,j,k}=\left| \log_{10} p_{i,j,k}-\log_{10} median(\mathbf{p}_{j,k}) \right|$$

Equation 1

where *i* represents an individual, *j* represents a channel, *k* represents an eye condition and **p** represents the SAGES sample of powers for each channel and condition. In both SAGES and INTUIT/PRIME Data Sets, the median is defined as the median of the entire SAGES Data Set for each electrode and condition. The MoCA scores were transformed using the z-score. The z-scores for both SAGES and INTUIT/PRIME Data Sets were computed using the sample mean and standard deviation of the SAGES Data Set. MoCA was used as a feature in both feature sets. Thus, in both feature sets tested, a total of 7 features were used: 6 EEG powers and 1 cognitive score (Supplementary Table S5). The feature set with the alpha powers and MoCA will be referred to throughout this paper as the Principal Feature Set (middle column of Supplementary Table S5). The feature set containing the subsets of the alpha powers and MoCA will be referred to as the Sub-Alpha Feature Set (right column of Supplementary Table S5). The performance of both feature sets was benchmarked against the performance of the models trained on MoCA alone, referred to as MoCA-Alone. Two other reference sets of features for comparison are Alpha Powers-Alone and Sub-Alpha Powers-Alone, which consist of only EEG features (no MoCA).

**Models**

The following models were tested during the model selection step: logistic regression with L2 regularization with inner cross-validation (CV) for hyperparameter tuning; linear discriminant analysis (LDA) with inner CV for hyperparameter tuning; linear discriminant analysis (LDA) using Ledoit-Wolf estimator (LDA LW); LDA using Oracle Shrinkage Approximation estimator (LDA OA); nearest shrunken centroid (NSC) using Manhattan distance metric; NSC using Euclidean distance metric; Gaussian Naïve Bayes (GNB) using empirical priors; GNB using a priori-defined class priors (80-20 ratio) and decision tree. When computing AUC of the ROC and PR curves, Gaussian Naïve Bayes using empirical priors, GNB using a priori defined class priors and decision tree used class probability to compute the threshold (*predict_proba* in scikit-learn). All of the other models used decision function to compute the threshold (*decision_function* in scikit-learn).

**Chance-Level**

The formulas for chance-level metrics for imbalanced 2-class sample are as followed:

Accuracy = prob_pos * samp_pos + prob_neg * samp_neg

Sensitivity = (prob_pos * samp_pos) / (prob_pos * samp_pos + prob_neg * samp_pos)

Specificity = (prob_neg * samp_neg) / (prob_pos * samp_neg + prob_neg * samp_neg)

AUC ROC = 0.5

F1 score = (2 * prob_pos * samp_pos) / (prob_pos + samp_pos)

AUC PR = samp_pos

PPV = (prob_pos * samp_pos) / (prob_pos * samp_pos + prob_pos * samp_neg)

NPV = (prob_neg * samp_neg) / (prob_neg * samp_pos + prob_neg * samp_neg)

where prob_pos is the probability of the label being positive, prob_neg is the probability of the label being negative, samp_pos is the proportion of the sample being positive, samp_neg is the proportion of the sample being negative.

**Results**

The cross-validated results using Sub-Alpha Powers + MoCA on SAGES Data Set are shown in Supplementary Figure S8 as well as in Table S6A. LDA CV model has the highest f1-score (mean ± 95% confidence interval: 0.593). LDA CV’s sensitivity and specificity are 0.58 ± 0.09 and 0.92 ± 0.02, respectively. When retraining LDA CV on the entire SAGES Data Set, the results tested on INTUIT/PRIME Data Set are shown in Figure S9 and Table S6B. Its f1-score is 0.60, 95% CI [0.48, 0.80]. The ROC and precision-recall curves are shown in Figure S7B and their AUCs are 0.95, 95% CI [0.87, 1.00] and 0.73, 95% CI [0.49, 0.98], respectively. When benchmarked against MoCA-Alone with the same training and test protocol (Table S7B), the Sub-Alpha Powers + MoCA has better performance in all metrics.

**Table S1**. Demographics of SAGES excluded cohort (n=5)

|  | No Delirium  (n=5) |
| --- | --- |
| Age, mean year (SD) | 73 (3.9) |
| Female, n (%) | 1 (20.0) |
| MoCA, mean score (SD) | 26.1 (1.7) |
| **Surgery type, n (%)** |  |
| Total knee replacement | 3 (60.0) |
| Total hip replacement | 1 (20.0) |
| Other | 1 (20.0) |
| **Anesthesia type, n (%)** |  |
| Spinal | 4 (80.0) |
| General | 1 (20.0) |
| General and Spinal | 0 (0.00) |

**Table S2**. Demographics of INTUIT/PRIME excluded cohort (n=11)

|  | Full Sample  (n=11) | Delirium  (n=1) | No Delirium  (n=10) |
| --- | --- | --- | --- |
| Age, mean year (SD) | 70.7 (7.8) | 81 (0.0) | 69.7 (7.5) |
| Female, n (%) | 8 (81.8) | 0 (0.0) | 8 (80.0) |
| MMSE, mean score (SD) | 27.6 (1.8) | 25 (0.0) | 27.9 (1.6) |
| MoCA, mean score (SD) | 23.7 (2.6) | 20 (0.0) | 24.1 (2.5) |
| **Surgery type, n (%)** |  |  |  |
| Total knee replacement | 1 (9.1) | 0 (0.0) | 1 (10.0) |
| Total hip replacement | 0 (0.0) | 0 (0.0) | 0 (0.0) |
| Other | 10 (90.9) | 1 (100.0) | 9 (90.0) |
| **Anesthesia type, n (%)** |  |  |  |
| General | 9 (81.8) | 1 (100.0) | 8 (80.0) |
| General and Neuraxial Block | 1 (9.1) | 0 (0.0) | 1 (10.0) |
| MAC/Neuraxial Block | 1 (9.1) | 0 (0.0) | 1 (10.0) |

**Table S3.** MMSE to MoCA Crosswalk.

(For details, see Saczynski et al^2^.)

| **MMSE** | **MoCA** |
| --- | --- |
| 30 | 28.5 |
| 29 | 25.5 |
| 28 | 24 |
| 27 | 22.5 |
| 26 | 21 |
| 25 | 20 |
| 24 | 18.5 |
| 23 | 17 |
| 22 | 16 |
| 21 | 14.5 |
| 20 | 13 |
| 19 | 12 |
| 18 | 11.5 |
| 17 | 9 |
| 16 | 8 |
| 15 | 7 |
| 14 | 6 |
| 12 | 5 |
| etc. |  |

**Table S4.** List of Frequency Bands and ROIs Explored

| **Frequency Bands** | **Ratio** | **ROIs** | **ROI channel list** |
| --- | --- | --- | --- |
| Delta [1, 4] Hz | (Alpha + Beta)/(Delta + Theta) | Eyes | F7, Fp1, Fp2, F8 |
| Theta [4, 8] Hz | (Alpha)/(Delta + Theta) | Frontal | F3, F4, Fz |
| Alpha [8, 12] Hz |  | Left Frontal | F3 |
| Beta [12, 20] Hz |  | Right Frontal | F4 |
|  |  | Left Frontal Temporal | F7, T7 |
| [4,6] Hz |  | Right Frontal Temporal | F8, T8 |
| [6, 8] Hz |  | Central | FC5, FC1, FC2, FC6, C3, Cz, C4, CP5, CP1, CP2, CP6 |
| [8, 10] Hz |  | Left Occipital | P3 |
| [10, 12] Hz |  | Right Occipital | P4 |
| [9, 11] Hz |  | Occipital | P3, Pz, P4, POz, O1, O2 |
|  |  | Left Parietal Temporal | P7 |
|  |  | Right Parietal Temporal | P8 |

**Table S5.** Cross-validation performances of SPR Feature Set, Frontal Alpha Feature Set and the Principal Feature Set (Alpha Powers + MoCA). Ranges enclosed in brackets represent 95% confidence intervals. Green highlighted row represents the model with the highest f1-score. Note: the f1-score is 0.574 for LDA LW and 0.565 for LDA OA.

| **Model Selection: SPR** | | | | | | | | |
| --- | --- | --- | --- | --- | --- | --- | --- | --- |
| Model Name | Accuracy | Sensitivity | Specificity | F1 | ROC AUC | PR AUC | PPV | NPV |
| Lasso L2 CV | 0.84  [0.83, 0.86] | 0.18  [0.12, 0.24] | 0.96  [0.94, 0.97] | 0.21  [0.14, 0.29] | 0.57  [0.51, 0.62] | 0.37  [0.32, 0.42] | 0.26  [0.16, 0.37] | 0.88  [0.87, 0.89] |
| LDA CV | 0.85  [0.83, 0.88] | 0.34  [0.25, 0.43] | 0.94  [0.92, 0.96] | 0.38  [0.29, 0.47] | 0.71  [0.66, 0.76] | 0.51  [0.45, 0.58] | 0.44  [0.32, 0.55] | 0.90  [0.88, 0.91] |
| LDA LW | 0.87  [0.86, 0.89] | 0.39  [0.30, 0.47] | 0.95  [0.94, 0.97] | 0.45  [0.37, 0.53] | 0.73  [0.68, 0.78] | 0.53  [0.47, 0.60] | 0.54  [0.43, 0.66] | 0.91  [0.89, 0.92] |
| LDA OA | 0.87  [0.85, 0.89] | 0.41  [0.34, 0.49] | 0.94  [0.93, 0.96] | 0.48  [0.40, 0.56] | 0.76  [0.71, 0.80] | 0.54  [0.48, 0.60] | 0.56  [0.45, 0.67] | 0.91  [0.89, 0.92] |
| NSC MH | 0.59  [0.56, 0.63] | 0.52  [0.43, 0.61] | 0.60  [0.57, 0.64] | 0.27  [0.22, 0.31] | 0.56  [0.51, 0.62] | 0.36  [0.30, 0.41] | 0.18  [0.15, 0.21] | 0.88  [0.86, 0.91] |
| NSC Custom EC | 0.80  [0.77, 0.83] | 0.53  [0.43, 0.62] | 0.85  [0.80, 0.89] | 0.44  [0.37, 0.51] | 0.72  [0.67, 0.78] | 0.57  [0.50, 0.64] | 0.38  [0.30, 0.46] | 0.92  [0.90, 0.93] |
| GNB | 0.68  [0.65, 0.71] | 0.19  [0.13, 0.25] | 0.76  [0.72, 0.79] | 0.15  [0.10, 0.20] | 0.48  [0.42, 0.54] | 0.24  [0.21, 0.27] | 0.12  [0.07, 0.18] | 0.85  [0.83, 0.86] |
| GNB Prior | 0.64  [0.60, 0.67] | 0.19  [0.12, 0.26] | 0.71  [0.66, 0.75] | 0.12  [0.08, 0.16] | 0.47  [0.41, 0.53] | 0.23  [0.19, 0.27] | 0.09  [0.06, 0.12] | 0.84  [0.82, 0.85] |
| Tree | 0.79  [0.76, 0.81] | 0.32  [0.24, 0.40] | 0.86  [0.84, 0.89] | 0.30  [0.23, 0.36] | 0.59  [0.55, 0.63] | 0.24  [0.20, 0.28] | 0.28  [0.20, 0.36] | 0.89  [0.87, 0.90] |
| **Model Selection: Frontal Alpha** | | | | | | | | |
| Model Name | Accuracy | Sensitivity | Specificity | F1 | ROC AUC | PR AUC | PPV | NPV |
| Lasso L2 CV | 0.90  [0.88, 0.91] | 0.36 [0.28, 0.44] | 0.98 [0.97, 0.99] | 0.46 [0.36, 0.55] | 0.73 [0.68, 0.79] | 0.60 [0.52, 0.67] | 0.61 [0.48, 0.74] | 0.90 [0.89, 0.92] |
| LDA CV | 0.85  [0.84, 0.87] | 0.40 [0.32, 0.48] | 0.93 [0.91, 0.94] | 0.42 [0.34, 0.50] | 0.73 [0.67, 0.79] | 0.59 [0.52, 0.67] | 0.44 [0.35, 0.54] | 0.91 [0.90, 0.92] |
| LDA LW | 0.86  [0.84, 0.87] | 0.43 [0.36, 0.50] | 0.93 [0.91, 0.95] | 0.47 [0.40, 0.53] | 0.77 [0.73, 0.81] | 0.65 [0.60, 0.70] | 0.51 [0.42, 0.60] | 0.91 [0.90, 0.92] |
| LDA OA | 0.85  [0.83, 0.87] | 0.41 [0.33, 0.49] | 0.92 [0.90, 0.94] | 0.44 [0.36, 0.52] | 0.74 [0.69, 0.80] | 0.60 [0.53, 0.68] | 0.47 [0.37, 0.58] | 0.91 [0.89, 0.92] |
| NSC MH | 0.70  [0.66, 0.74] | 0.34 [0.25, 0.43] | 0.76 [0.71, 0.81] | 0.27 [0.21, 0.34] | 0.60 [0.54, 0.66] | 0.38 [0.32, 0.44] | 0.23 [0.16, 0.30] | 0.88 [0.86, 0.89] |
| NSC Custom EC | 0.82  [0.79, 0.84] | 0.47 [0.39, 0.56] | 0.87 [0.84, 0.90] | 0.42 [0.35, 0.48] | 0.71 [0.66, 0.76] | 0.52 [0.45, 0.59] | 0.37 [0.29, 0.46] | 0.92 [0.90, 0.93] |
| GNB | 0.48  [0.44, 0.51] | 0.69 [0.60, 0.78] | 0.45 [0.41, 0.48] | 0.27 [0.24, 0.30] | 0.73 [0.67, 0.79] | 0.60 [0.52, 0.68] | 0.17 [0.14, 0.19] | 0.90 [0.87, 0.93] |
| GNB Prior | 0.46  [0.43, 0.49] | 0.72 [0.63, 0.81] | 0.42 [0.38, 0.46] | 0.28 [0.24, 0.31] | 0.70 [0.65, 0.76] | 0.58 [0.50, 0.66] | 0.17 [0.15, 0.19] | 0.91 [0.88, 0.94] |
| Tree | 0.79  [0.77, 0.82] | 0.42 [0.34, 0.50] | 0.86 [0.83, 0.88] | 0.36 [0.30, 0.43] | 0.64 [0.60, 0.67] | 0.26 [0.22, 0.30] | 0.32 [0.25, 0.39] | 0.90 [0.89, 0.92] |
| **Model Selection: Principal Feature Set** | | | | | | | | |
| Model Name | Accuracy | Sensitivity | Specificity | F1 | ROC AUC | PR AUC | PPV | NPV |
| Lasso L2 CV | 0.89  [0.87, 0.90] | 0.37  [0.30, 0.45] | 0.97  [0.96, 0.98] | 0.46  [0.38, 0.55] | 0.82  [0.78, 0.86] | 0.66  [0.59, 0.72] | 0.61  [0.49, 0.73] | 0.91  [0.89, 0.92] |
| LDA CV | 0.87  [0.85, 0.89] | 0.59  [0.49, 0.69] | 0.92  [0.90, 0.94] | 0.56  [0.47, 0.64] | 0.79  [0.74, 0.84] | 0.67  [0.60, 0.74] | 0.52  [0.43, 0.62] | 0.93  [0.92, 0.95] |
| LDA LW | 0.88  [0.86, 0.90] | 0.54  [0.45, 0.62] | 0.94  [0.93, 0.96] | 0.57  [0.50, 0.65] | 0.80  [0.76, 0.85] | 0.66  [0.59, 0.72] | 0.62  [0.52, 0.71] | 0.93  [0.91, 0.94] |
| LDA OA | 0.87  [0.85, 0.89] | 0.52  [0.44, 0.60] | 0.93  [0.91, 0.95] | 0.57  [0.50, 0.63] | 0.80  [0.75, 0.84] | 0.67  [0.62, 0.73] | 0.61  [0.52, 0.71] | 0.92  [0.91, 0.94] |
| NSC MH | 0.79  [0.76, 0.82] | 0.60  [0.50, 0.70] | 0.82  [0.78, 0.86] | 0.47  [0.41, 0.54] | 0.80  [0.77, 0.84] | 0.51  [0.45, 0.57] | 0.39  [0.32, 0.47] | 0.93  [0.92, 0.95] |
| NSC Custom EC | 0.83  [0.81, 0.85] | 0.58  [0.50, 0.66] | 0.87  [0.84, 0.89] | 0.50  [0.44, 0.55] | 0.84  [0.80, 0.88] | 0.65  [0.59, 0.71] | 0.43  [0.37, 0.50] | 0.93  [0.92, 0.94] |
| GNB | 0.78  [0.76, 0.81] | 0.74  [0.67, 0.81] | 0.79  [0.76, 0.82] | 0.51  [0.46, 0.56] | 0.89  [0.86, 0.92] | 0.72  [0.66, 0.78] | 0.39  [0.34, 0.44] | 0.95  [0.94, 0.96] |
| GNB Prior | 0.77  [0.74, 0.80] | 0.81  [0.74, 0.88] | 0.76  [0.73, 0.80] | 0.53  [0.49, 0.57] | 0.88  [0.85, 0.92] | 0.71  [0.65, 0.77] | 0.39  [0.35, 0.44] | 0.97  [0.95, 0.98] |
| Tree | 0.80  [0.78, 0.82] | 0.40  [0.32, 0.49] | 0.87  [0.85, 0.89] | 0.35  [0.28, 0.42] | 0.64  [0.60, 0.68] | 0.27  [0.22, 0.31] | 0.31  [0.24, 0.38] | 0.90  [0.89, 0.91] |

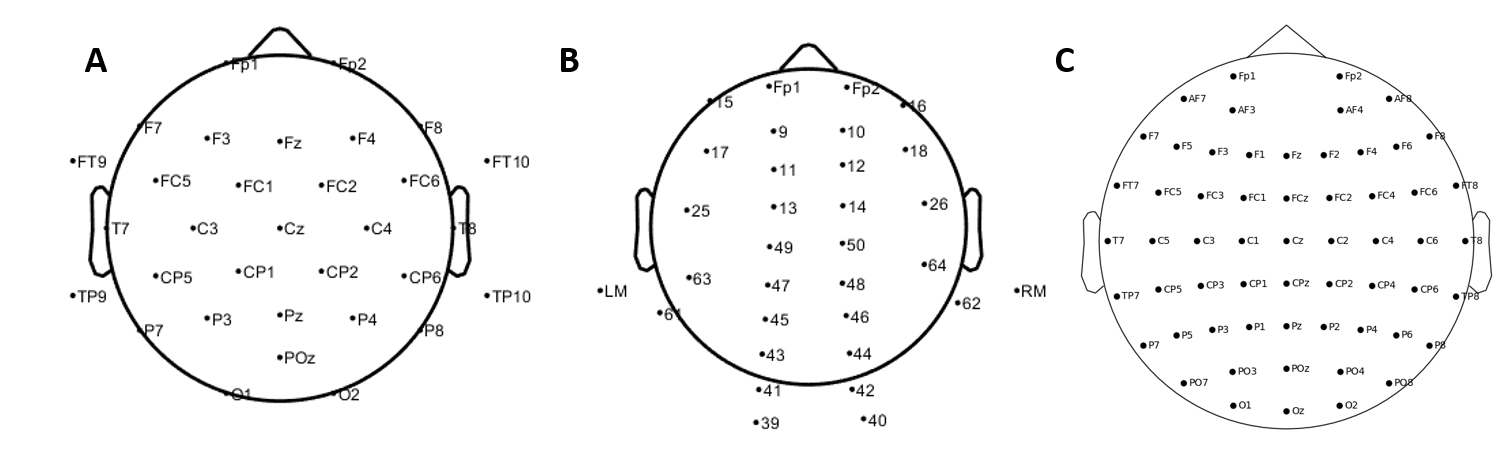

**Supplementary Figure S1.** A) Standard international 32-channel “10-20” EEG Montage used in a sub-cohort of INTUIT/PRIME study. Please note it’s not a true 10-20, but just named “10-20”. B) Custom 32-channel EEG Montage used in a sub-cohort of INTUIT/PRIME study. C) 64-channel according to 10-20 standard international system used in SAGES study.

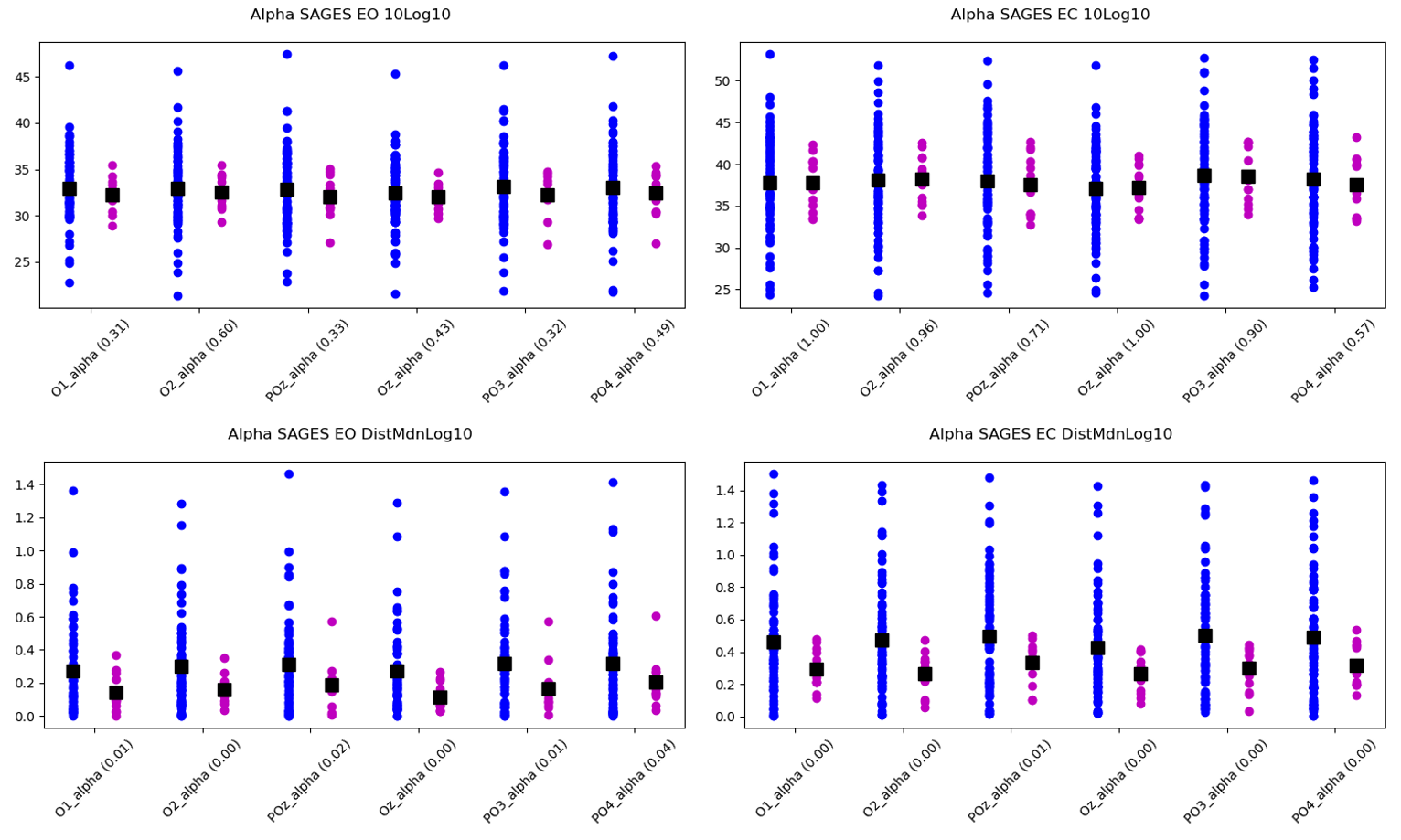

**Supplementary Figure S2** Scatter plots of the alpha powers in the SAGES Data Set. Left column shows EO condition, right column shows EC condition. Top row shows untransformed (10*log10 of alpha power) and bottom row shows transformed alpha powers (arbitrary unit).

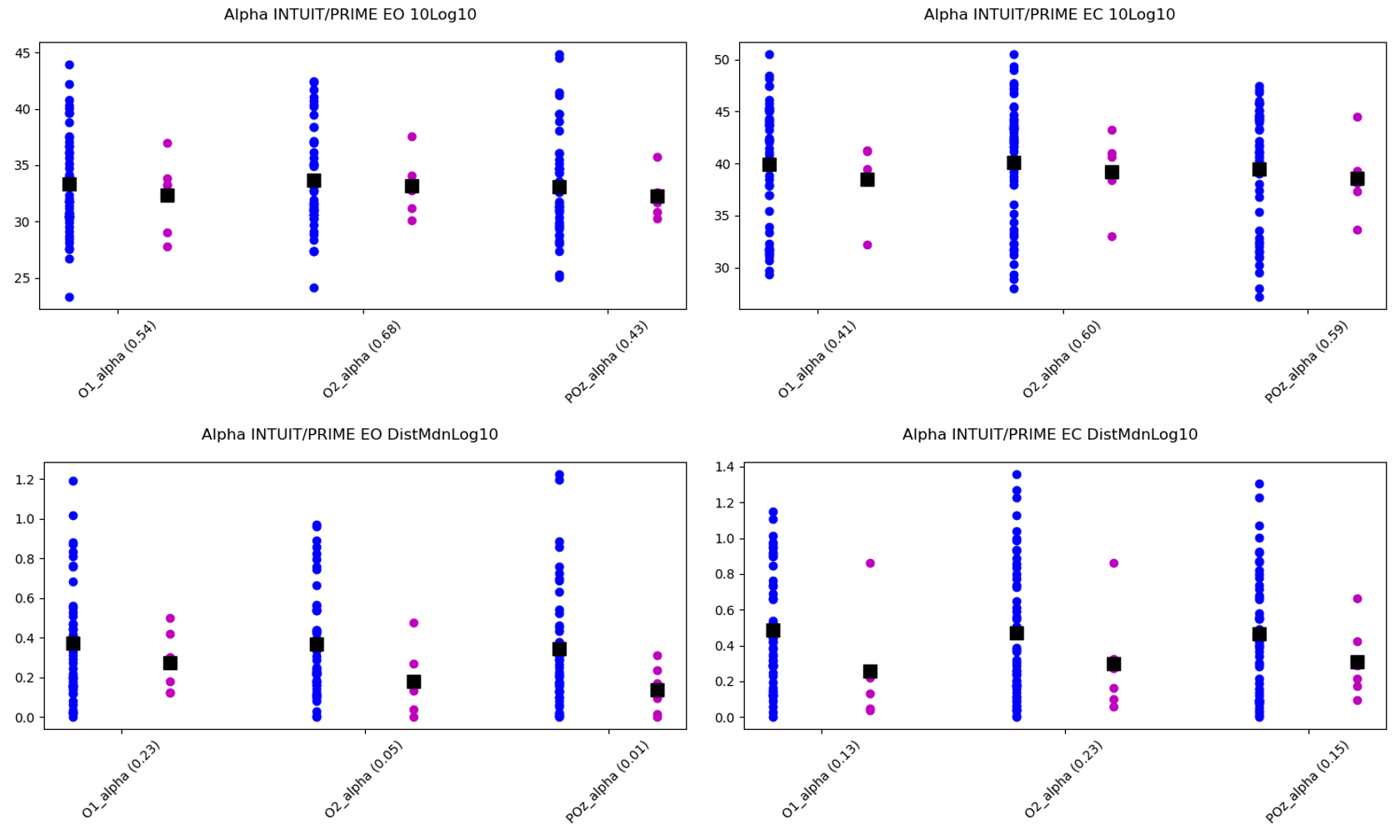

**Supplementary Figure S3** Same layout as Supplementary Figure S2 but for the INTUIT/PRIME Data Set.

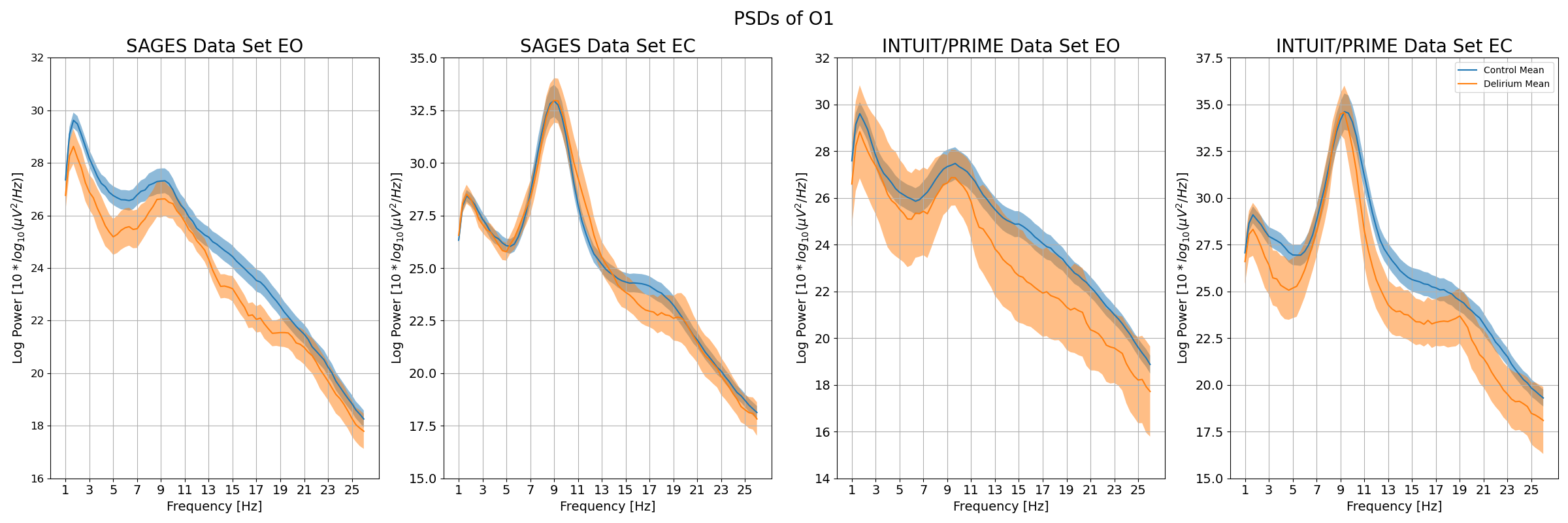

**Supplementary Figure S4.** PSDs of O1 channel from both SAGES and INTUIT/PRIME Data Sets in both EO and EC conditions.

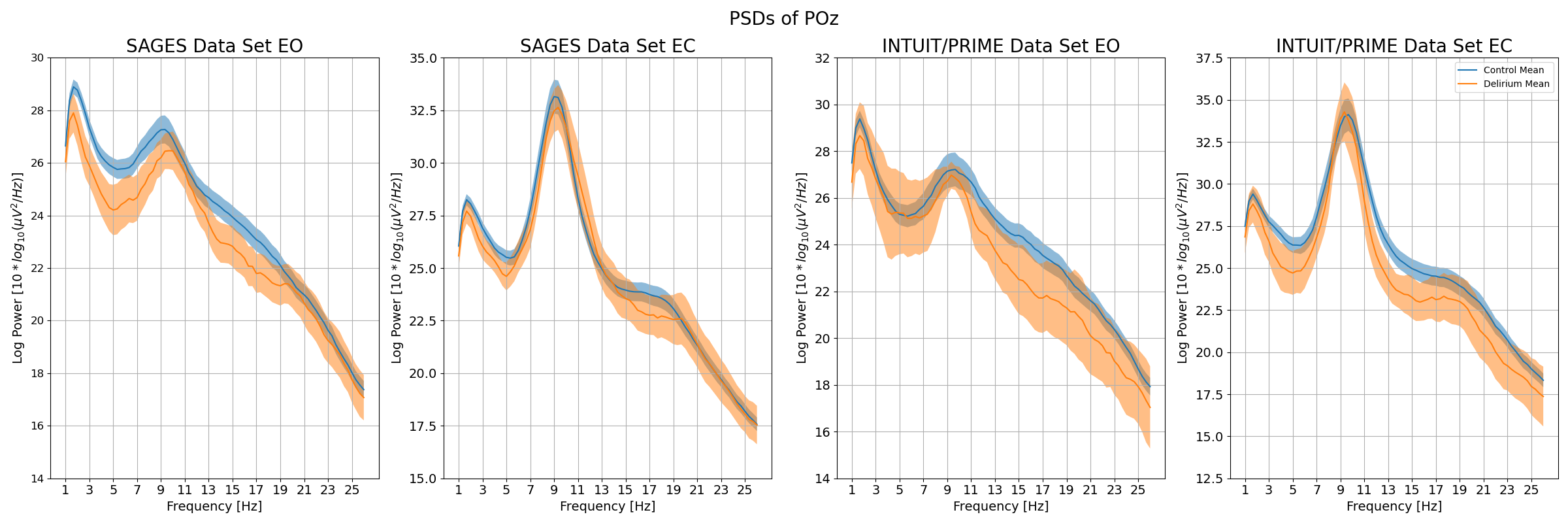

**Supplementary Figure S5.** PSDs of POz channel from both SAGES and INTUIT/PRIME Data Sets in both EO and EC conditions.

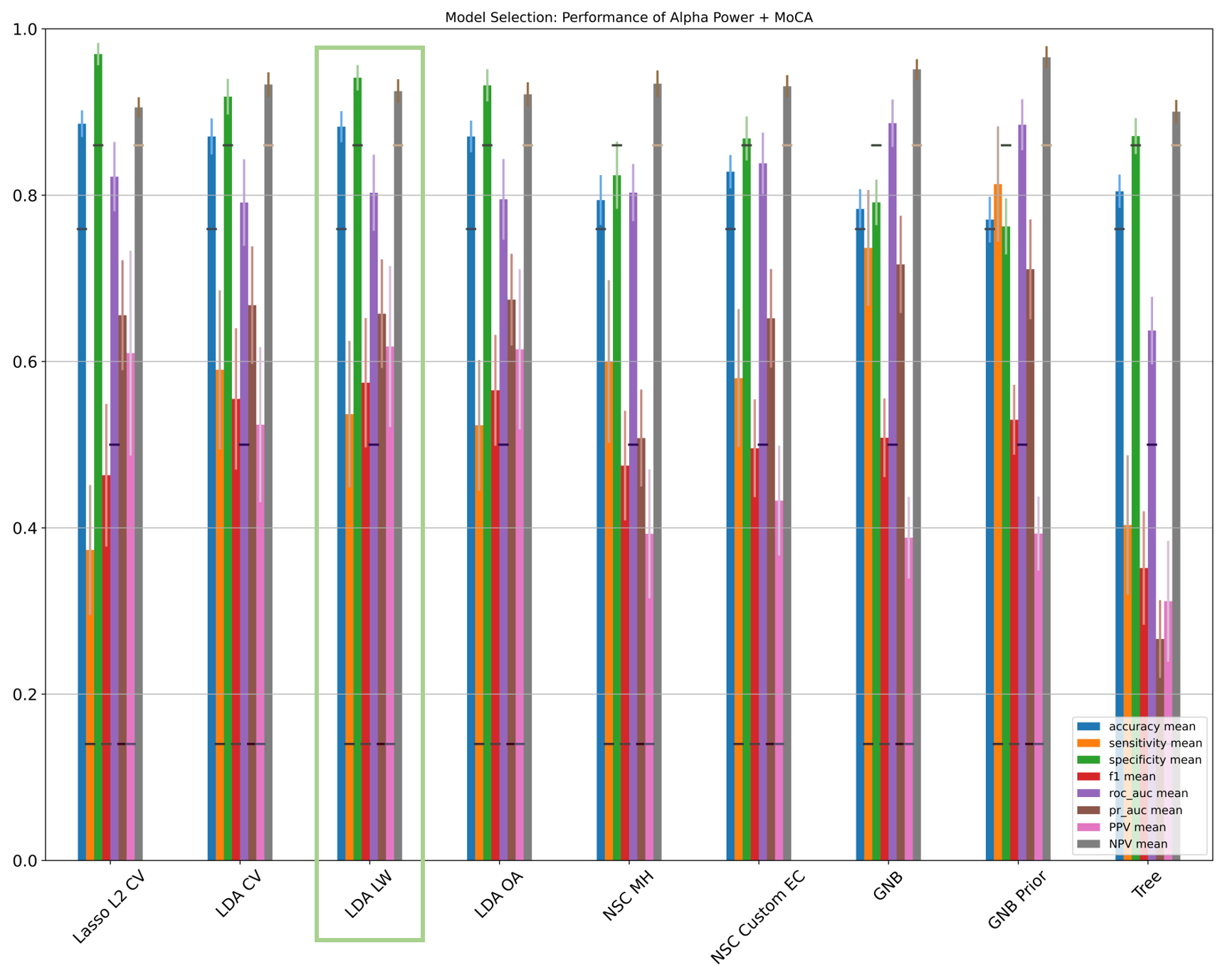

Supplementary Figure S6

1. Helfand BKI, Detroyer E, Milisen K, et al. Harmonization of Four Delirium Instruments: Creating Crosswalks and the Delirium Item-Bank (DEL-IB). The American Journal of Geriatric Psychiatry. 2022;30(3):284-294. doi:10.1016/j.jagp.2021.07.011
2. Saczynski, J. S. *et al.* The Montreal Cognitive Assessment: Creating a Crosswalk with the Mini‐Mental State Examination. *J American Geriatrics Society* **63**, 2370–2374 (2015).
