## Supplementary material for "Prediction of Postoperative Delirium in Older Adults from Preoperative Cognition and Occipital Alpha Power from Resting-State Electroencephalogram": Table_01

|  | Full sample | Delirium | No Delirium | p-value (t-statistic) |
| --- | --- | --- | --- | --- |
|  | (n=85) | (n=12) | (n=73) | (dof=83) |
| Age, mean year (SD) | 73 (6.4) | 75 (9.5) | 72 (5.8) | -- |
| Female, n (%) | 56 (65.9) | 10 (83.3) | 46 (63.0) | -- |
| MoCA, mean score (SD) | 26.1 (3.4) | 21.8 (5.4) | 26.8 (2.4) | 2.09e-6 (5.10) |
| **Surgery type, n (%)** |  |  |  |  |
| Total knee replacement | 52 (61.2) | 8 (66.7) | 44 (60.3) | -- |
| Total hip replacement | 25 (29.4) | 3 (25.0) | 22 (30.1) | -- |
| Other | 8 (9.4) | 1 (8.3) | 7 (9.6) | -- |
| **Anesthesia type, n (%)** |  |  |  |  |
| Spinal | 78 (91.8) | 11 (91.7) | 67 (91.8) | -- |
| General | 7 (8.2) | 1 (8.3) | 6 (8.2) | -- |
| General and Spinal | 0 (0.0) | 0 (0.0) | 0 (0.0) | -- |

Table 1 Demographics of SAGES analysis sample. Two-tailed two-sample t-test was used to compute the p-value only for the MoCA scores.
