## Supplementary material for "Prediction of Postoperative Delirium in Older Adults from Preoperative Cognition and Occipital Alpha Power from Resting-State Electroencephalogram": Table_02

|  | Full sample | Delirium | No Delirium | p-value (t-statistic) |
| --- | --- | --- | --- | --- |
|  | (n=51) | (n=6) | (n=45) | (dof=49) |
| Age, mean year (SD) | 68.3 (5.2) | 71.2 (5.7) | 68.0 (5.2) | -- |
| Female, n (%) | 25 (49.0) | 1 (16.7) | 24 (53.3) | -- |
| MMSE, mean score (SD) | 27.5 (2.4) | 24.0 (2.6) | 28.0 (1.9) | 4.15e-5 (4.50) |
| MoCA Converted, mean score (SD) | 23.7 (3.6) | 18.5 (3.4) | 24.4 (3.0) | 8.82e-5 (4.27) |
| **Surgery type, n (%)** |  |  |  |  |
| Total knee replacement | 3 (5.9) | 0 (0.0) | 3 (6.7) | -- |
| Total hip replacement | 1 (2.0) | 0 (0.0) | 1 (2.2) | -- |
| Other | 47 (92.1) | 6 (100.0) | 41 (91.1) | -- |
| **Anesthesia type, n (%)** |  |  |  |  |
| General | 39 (76.5) | 4 (66.7) | 35 (77.8) | -- |
| General and Neuraxial Block | 8 (15.7) | 2 (33.3) | 6 (13.3) | -- |
| MAC/Neuraxial Block | 3 (5.9) | 0 (0.0) | 3 (6.7) | -- |

Table 2 Demographics of INTUIT/PRIME analysis sample. Two-tailed two-sample t-test was used to compute the p-value only for the MMSE and MoCA Converted scores.
