## Supplementary material for "Prediction of Postoperative Delirium in Older Adults from Preoperative Cognition and Occipital Alpha Power from Resting-State Electroencephalogram": Table_03

**Table 3.** Performances of A) Principal Feature Set (Alpha-Powers + MoCA), B) MoCA-Alone and C) Alpha-Powers-Alone on INTUIT/PRIME Data Set. Ranges enclosed in brackets represent 95% confidence intervals.

| A) **Principal Feature Set (Alpha-Powers + MoCA)** | | | | | | | | |
| --- | --- | --- | --- | --- | --- | --- | --- | --- |
|  | Accuracy | Sensitivity | Specificity | F1 | ROC AUC | PR AUC | PPV | NPV |
| LDA LW | 0.90  [0.80, 0.98] | 0.83  [0.50, 1.00] | 0.91  [0.82, 0.98] | 0.67  [0.43, 0.91] | 0.94  [0.86, 0.99] | 0.70  [0.44, 0.95] | 0.56  [0.33, 0.86] | 0.98  [0.93, 1.00] |
| B) **MoCA Alone** | | | | | | | | |
|  | Accuracy | Sensitivity | Specificity | F1 | ROC AUC | PR AUC | PPV | NPV |
| LDA LW | 0.80  [0.69, 0.90] | 0.83  [0.50, 1.00] | 0.80  [0.67, 0.91] | 0.50  [0.31, 0.71] | 0.91  [0.81, 0.98] | 0.54  [0.30, 0.88] | 0.36  [0.21, 0.56] | 0.97  [0.92, 1.00] |
| C) **Alpha-Powers Alone** | | | | | | | | |
|  | Accuracy | Sensitivity | Specificity | F1 | ROC AUC | PR AUC | PPV | NPV |
| LDA LW | 0.90  [0.88, 0.94] | 0.17  [0.00, 50] | 1.00  [1.00, 1.00] | 0.29  [0.00, 0.67] | 0.77  [0.57, 0.94] | 0.53  [0.21, 0.87] | 1.00  [0.00, 1.00] | 0.90  [0.88, 0.94] |
